# Assessing the Long-Term Economic Impact of Wheezing Episodes After Severe RSV Disease in Children from Argentina: A Cost of Illness Analysis

**DOI:** 10.1101/2024.03.18.24304483

**Authors:** Julia Dvorkin, Clint Pecenka, Emiliano Sosa, Andrea Sancilio, Karina Dueñas, Andrea Rodriguez, Carlos Rojas-Roque, Patricia B. Carruitero, Ranju Baral, Elisabeth Vodicka, Fernando P. Polack, Romina Libster, Mauricio T. Caballero

## Abstract

**Introduction:** There is a lack of available data on the economic burden of wheezing episodes resulting from prior severe respiratory syncytial virus (RSV) infections in resource-constrained settings. This study aimed to assess the cost incurred for wheezing episodes during five years after a severe RSV infection in children from Argentina, considering both the public health system and societal perspectives.

**Methods:** A prospective cohort was conducted to assess the cost-of-illness (COI) linked to wheezing episodes after severe RSV disease in children from Buenos Aires, Argentina. Direct medical and non-medical costs were estimated, along with indirect costs per episode and patient. Data pertaining to healthcare resource utilization, indirect expenses, and parental out-of-pocket costs were obtained from research forms. The overall cost per hospitalization and health visits were calculated from the perspectives of the healthcare system and society. Costs were quantified in US dollars.

**Results:** Overall, 150 children aged between 12 and 60 months presented a total of 429 wheezing episodes. The median number of wheezing episodes per patient was 5 (IQR 3-7). The mean cost per wheezing episode was US$ 191.01 (95% confidence interval [CI] $166.37 – $215.64). The total cost per episode of wheezing was significantly higher (p<0.001) in infants under 12 months of age (207.43, 95%CI 154.3-260.6) compared to older toddler subgroups. The average cumulative cost associated to wheezing per patient was US$ 415.99 (95%CI $313.35 - $518.63). Considering both acute RSV disease and long-term wheezing outcomes the cumulative mean cost per patient was US$ 959.56 (95%CI $832.01-$1087.10).

**Conclusions:** This study reveals the economic impact of prolonged wheezing resulting from severe acute RSV infection on Argentina’s public health system and society. The estimates obtained serve as valuable inputs for informing cost-effectiveness analyses of upcoming RSV preventive interventions.

**What is already known on this topic:** Multiple studies demonstrate the association between severe acute lower respiratory tract infections caused by RSV in infancy with long-term obstructive pulmonary disease such as recurrent wheezing or asthma. Nevertheless, there is a lack of information regarding the economic impact of these frequent wheezing episodes in individuals who experienced hospitalization due to RSV disease early in life, particularly in low- and middle-income countries. To address this gap, we conducted a prospective cohort study to ascertain the cost of illness associated with wheezing episodes in children during their initial 5 years of life following a severe RSV infection within a low-income population in Buenos Aires, Argentina.

**What this study adds:** This study provides a comprehensive account of both medical and non-medical expenses associated with frequent wheezing episodes in childhood in low settings of Argentina, focusing on patients who experienced a severe RSV infection. Furthermore, we computed the total cost, encompassing the expenses associated with the initial severe RSV disease in those patients with long-term wheezing episodes.

**How this study might affect research, practice or policy:** The data produced in this study is important for estimating the economic impact of forthcoming preventive measures against RSV in low- and middle-income countries through cost-effectiveness studies. Health decision-makers can leverage this information for future decisions on implementing preventive policies against RSV in infancy.

## Introduction

Respiratory Syncytial Virus (RSV) is a ubiquitous pathogen that poses a considerable threat to infants and young children globally, causing millions of hospitalizations due to acute lung infection and more than 100,000 deaths every year^1–4^. More than ninety percent of the fatal cases associated with this virus occur in contrasting settings of low- and middle-income countries (LMICs)^5^. In Argentina, the mortality rate for RSV is high, reaching more than 0.5 per 1,000 live births, with more than fifty percent of the fatal cases taking place at home in the absence of medical attention^6–9^.

While the immediate clinical impact of severe RSV infections is well-documented, there is an emerging recognition of the long-term sequelae that can follow an early and severe acute lower respiratory tract illness (ALRTI) caused by RSV, particularly the development of recurrent wheezing and asthma^10–15^. Recurrent wheezing, characterized by episodic wheezing and breathing difficulties, can persist for years after an initial severe RSV infection and often requires ongoing medical care and treatment^16,17^. These repeated episodes not only affect the quality of life for children and their families but also impose a substantial economic burden on the healthcare system and society as a whole^18,19^. In the United States, the combined financial expenses associated with various types of asthma amount to US$14 billion, comprising US$9.4 billion in direct expenditures and US$4.6 billion in indirect costs, which encompass missed school and workdays^20^. Costs related to recurrent wheezing in preschool children are estimated to range widely from US$1,020 to US$29,000 per child per year in US and Canada^21,22^. However, the economic impact of recurrent wheezing as an association of severe RSV disease, has not yet been extensively explored in LMIC. While some research has been conducted on the clinical aspects of RSV infections and their associated costs in Argentina and other LMICs^23,24^, there is a considerable lack of comprehensive assessments regarding the long-term effects of RSV infection on healthcare resource utilization and associated costs^25,26^.

Given the possibility of upcoming preventive measures, it is imperative to estimate the cost of illness (COI) for long-term wheezing outcomes associated to a previous severe ALRTI due to RSV in LMIC^4^. This is of utmost importance for several compelling reasons. To begin with, the majority of severe cases and fatalities occur within these nations, necessitating the swift deployment of preventive strategies to reduce the impact of RSV-related illnesses^13,27^. Additionally, studies on the COI offer valuable insights into the financial strain caused by RSV, thereby revealing potential cost savings in the absence of this disease. Lastly, COI investigations contribute to a more holistic comprehension of the condition, encompassing its economic implications^27^.

The main objective of this study is to examine the COI associated with long-term wheezing episodes in children who have experienced severe RSV infections in Argentina during their first year of life. By shedding light on the economic dimensions of this complex issue, we aim to inform healthcare policy makers, promote effective preventive strategies, and improve the overall well-being of affected children and their families.

## Methods

### Study design

We conducted a prospective cohort study for the evaluation of the COI associated with wheezing episodes that followed a severe RSV-ALRTI within the first year of life in children attended in two public hospitals of Buenos Aires, Argentina. The main study involved a prospective follow-up for 5 years of a cohort of patients under 12 months of age who were hospitalized due to RSV-ALRTI, allowing for a comprehensive assessment of the long-term economic impact of wheezing syndrome in this population. The analysis was carried out from both public health system and societal perspectives^28^. This study adhered to the methodological considerations previously published for studies on the COI^29,30^. Patients and the public were not involved in the design, conducting, reporting or dissemination plans of this study.

### Data source and study setting

This study employed data derived from a prospective, cohort established at two public hospitals located in the southern region of Buenos Aires, Argentina. The enrollment process of this cohort took place from May 2014 to August 2016, and the follow-up was concluded until June 2022. The cohort’s database encompasses details regarding the utilization of healthcare resources, RSV diagnoses, and demographic information of individuals who underwent ALRTI as previously described and detailed below in the instruments section and supplementary materials^26^.

### Instruments

For the collection of healthcare resources, data collection instruments were developed in REDCap as previously described^26,31^. Parents or responsible adults were subject to multiple interviews conducted via telephone calls to gather information regarding their employment status, monthly income, and itemized out-of-pocket expenses using research forms and uploading them to REDCap. For long-term wheezing outcomes, the study physicians filled out the research forms on each occasion when patients experienced respiratory exacerbation episodes, attended outpatient health clinics, or inpatient pediatric wards. In instances where health visits did not occur, study members maintained periodic contact (every 2 months) with the families through phone calls, WhatsApp messages, or email.

### Population

Infants aged 12 months or younger hospitalized with severe ALTRI were screened for eligibility. A comprehensive panel of 10 respiratory pathogens, including RSV, was identified in nasopharyngeal aspirates using quantitative real-time polymerase chain reaction (qPCR) in our research laboratory for each eligible patient^6,9^. Those infants diagnosed with of RSV by qPCR were included in this study^26^. Severe ALRTI was defined as the presence of at least one manifestation of lower respiratory tract infection sign (cough, nasal flaring, indrawing of the lower chest wall, subcostal retractions, stridor, rales, rhonchi, wheezing, crackles or crepitations, or observed apnea) plus hypoxemia (peripheral oxygen saturation of <95% at room air) or tachypnea (≥70 breaths per minute from 0 to 59 days of age, and ≥60 breaths per minute at 60 days of age or older)^6,32,33^.

Of the initially 470 patients screened for eligibility, 256 were confirmed to have severe RSV-ALRTI and enrolled in the study. They were then tracked for a minimum of 5 years to monitor the presence of wheezing episodes (see Supplementary Figure 1). The assessment of wheezing episodes was overseen by a study physician or periodically reported by parents through phone calls or face-to-face interviews using the ISAAC questionnaire^13,34^. Inclusion in the cost estimation required the presence of at least one documented episode of wheezing diagnosed by a physician following a severe RSV-ALRTI.

Loss of follow-up was deemed to occur if patients were unreachable through phone, email, or WhatsApp, did not visit the primary care center in the participant hospitals, or relocated to another town outside the designated catchment area of the study (Metropolitan Area of Buenos Aires).

### Outcome of the study

The study’s objective was to determine the overall cost, considering both direct and indirect costs, associated with hospitalizations and healthcare visits due to wheezing episodes^26^. All costs in this study were initially calculated in Argentinian pesos (ARS) at the time of data collection were obtained and subsequently converted to US dollars using the average exchange rate reported by the Central Bank of Argentina on that specific date^35^.

### Cost estimation

Medical costs estimation was based on the bottom-up costing method described elsewhere^26^. In this approach, healthcare resources used were meticulously identified and quantified through the data collection instrument. The unit cost associated with each healthcare resource was extracted from the financial database and the account records of the two participating hospitals^26^.

Through medical visits or phone calls, all 256 patients were followed up for at least five years and every wheezing episode was recorded along with the following information:

- Number of visits to the clinician per episode until recovery
- Severity of the episode according to the Pulmonary Index Score (only for medical visits)^36,37^: in every visit, presence of wheezing, accessory muscle use, rales and oxygen saturation were documented and then the episode classified under mild, moderate or severe ALRTI.
- Need for hospitalization.

For outpatient episodes, if the severity of illness in the visit to the clinician was moderate, a standard treatment on the emergency room was established that included the administration of albuterol and supplementary oxygen with a Venturi mask, systemic corticosteroids, and antipyretic drugs if necessary. The consultation fees for physicians and nurses were also considered in the total cost for outpatients.

For both outpatient and inpatient episodes, the total cost was disaggregated in the following components as previously described^26^: laboratory tests, labor cost, drugs, feeding, imaging diagnosis, supplies, oxygen supply, overhead, and equipment depreciation^26^. All these components costs were allocated as previously reported^26^. The total cost was calculated per episode and per patient, considering that every subject included may have had one or more episodes of wheezing (with or without hospitalization requirement).

To estimate the out-of-pocket expenditures, a two-part model was employed to address the substantial proportion of observations at zero combined with skewed positive outcomes (for non-zero)^38^. A similar approach has been used in a previous study^39^. In the first part, three logit models were used to estimate the probability of incurring out-of-pocket expenditures for meals, commute, and loss of income, respectively. Briefly, we modeled the natural log odds as a linear function of demographic (sex, age in months, patient’s weight, household income, parent’s education) and clinical (days of hospitalization, number of wheezing episodes) covariates. Mathematically, we estimated the following equation:

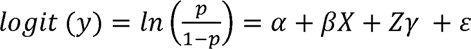

Where *p* is the probability of the outcome of interest *y*; *X* and *Z* are a subset of sociodemographic and clinical covariates; and *E* is the error term with distribution *E ∼ N*(0,1)^38^.

In the second part, a generalized linear model (GLM) with an identity link function and a gamma distribution for error terms was used to estimate the level of out-of-pocket expenditures for meals, commute, and loss of income. This model was chosen instead of the conventional ordinary least squares (OLS) regression because cost data often exhibit significant skewness and adhere to a non-normal distribution, potentially leading to violations of the fundamental assumptions of OLS^40,41^. The covariates included in the model were sociodemographic (sex, age in months, patient’s weight, household income, parent’s education) and clinical (days of hospitalization, number of wheezing episodes).

Lastly, the two parts were combined to estimate expected out-of-pocket expenditures by exploiting basic rule of probability^39^:

### Sample size and statistical analyses

Sample size was calculated using the following formula^42^:

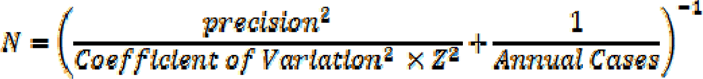

Precision was established at ±10% of the mean cost, assuming a coefficient of variation of 0.5, a z-score of 1.96 and p (wheezing episodes after severe RSV) of 62%. The minimum sample size necessary for assessing the costs of RSV-associated long-term wheeze was determined to be 115 participants.

We analyzed data using Stata (16.1, StataCorp LLC, College Station, TX). Descriptive statistics (frequency and percent) were used to summarize demographics, clinical variables, and healthcare resource utilization. We applied for the K-S test (Kolmogorov-Smirnov) to assess the normality of data. For normally distributed variables, we described data using means and standard deviations (SD); while we described data using the median and interquartile range (IQR) for non-normal distributed variables. We described the cost of recurrent wheezing episodes by using mean together with its 95% confidence interval (CI). We used Student’s T-test to compare average cost for outpatients and inpatients, and ANOVA to compare the mean direct medical cost of a wheezing episode between different age groups based on reported wheezing frequency rates (0-12 months, 12-24 months and >24 months of age)^11^.

### Ethical considerations

The institutional review boards at each participating hospital, the Province of Buenos Aires, and Vanderbilt University approved the study^26^. Informed consent was obtained from all participating parents or guardians.

## Results

### Study population

A total of 256 infants, all aged 12 months or less, hospitalized due to RSV-ALRTI, were included in this study, and followed for at least 5 years. The mean duration of follow-up for the 256 patients was 48.23 months (95% CI 44.50 to 51.99 months). Among them, 150 infants (58.6%) experienced at least one episode of wheezing after a severe ALRTI caused by RSV and were included in this analysis. The median number of wheezing episodes per patient reported during the follow-up was 5 (IQR 3-7, range 2- 11). The majority of the episodes occurred within two years after the first severe RSV-ALRTI: the median time until the last episode of wheezing was recorded is 22 months (IQR 12 - 33.5).

We compared sociodemographic and clinical-biological variables between patients who developed repeated episodes of wheezing during the first 5 years of life and those who did not (Table 1). Although both groups have similar characteristics, patients with repeated episodes of wheezing less frequently lived in precarious houses (built mostly with tin, wood, and/or mud) compared to controls (p=0.008). Across the population the predominant employment status among parents was informal work, with a substantial monthly income variability. The study determined an average household income of US$632.8 per month, leaving more than 70% of the families situated below the poverty line according to Argentina’s income thresholds per family of 4 members^43^.

**Table 1.**
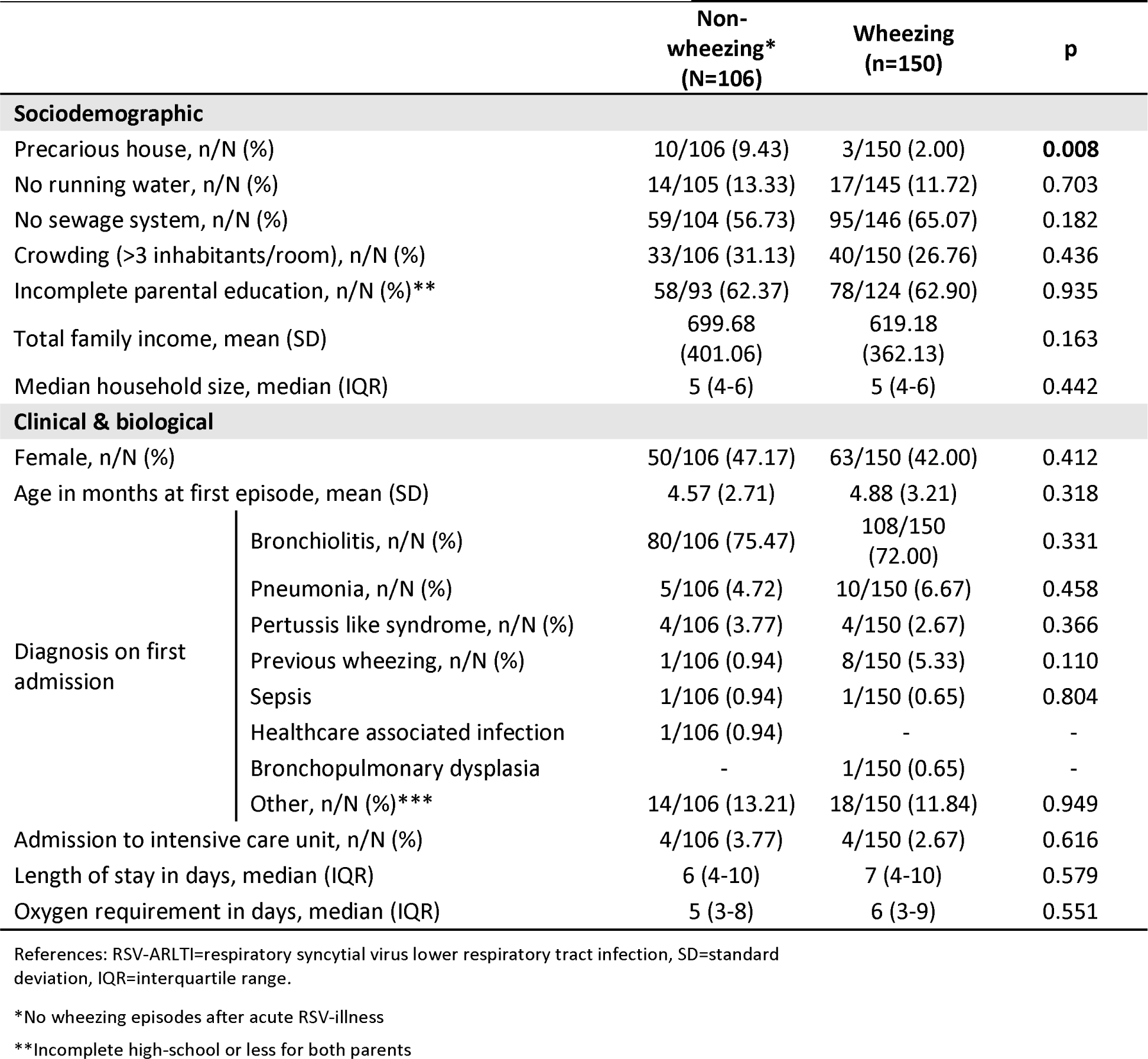
Characteristic of the study population.

Overall, from the total of 429 episodes of respiratory exacerbations with wheezing, 324 episodes received outpatient treatment, either through visits to the emergency room (ER) or pediatric primary care centers, and 105 episodes needed hospitalization in pediatric ward. A single wheezing episode may involve multiple health visits throughout its occurrence. No admission to pediatric intensive care units or fatal events related to episodic wheezing were recorded in this study.

### Direct medical costs of wheezing episodes

Considering both outpatient visits and hospitalizations, the mean cost per episode (including all health visits) was calculated at US$185.00 (95% CI $160.33 -$209.67). The mean direct medical cost for a severe wheezing episode necessitating hospitalization, including all health visits before hospital admission, was US$528.32 (95% CI $454.01 - $602.64), as detailed in Table 2. The aggregate cost for 105 hospital admissions due to wheezing episodes in 73 patients, requiring 716 hospital bed days (with a median stay of 5 days and an interquartile range of 3-8 days per episode), was US$60,729.81. In contrast, the average direct medical cost for a single episode not requiring hospital admission was US$85.92 (95% CI $80.51 - $91.34).

**Table 2.**
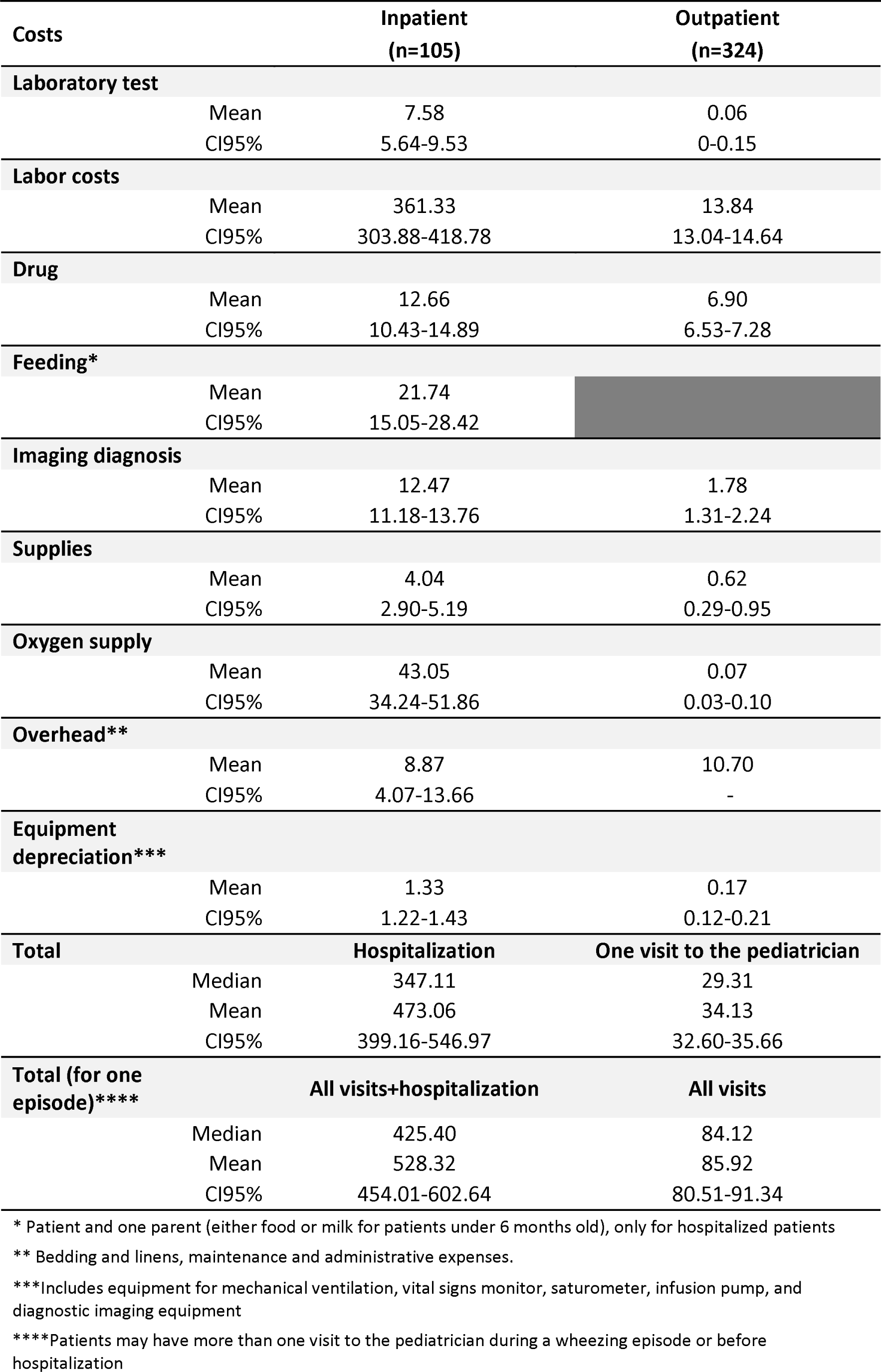
Comparative analysis of itemized costs between inpatient and outpatient consultations for wheezing episodes.

| Costs |  | Inpatient<br>(n=105) | Outpatient<br>(n=324) |
| --- | --- | --- | --- |
| **Laboratory test** |  |  |  |
|  | Mean | 7.58 | 0.06 |
|  | CI95% | 5.64-9.53 | 0-0.15 |
| **Labor costs** |  |  |  |
|  | Mean | 361.33 | 13.84 |
|  | CI95% | 303.88-418.78 | 13.04-14.64 |
| **Drug** |  |  |  |
|  | Mean | 12.66 | 6.90 |
|  | CI95% | 10.43-14.89 | 6.53-7.28 |
| **Feeding\*** |  |  |  |
|  | Mean | 21.74 |  |
|  | CI95% | 15.05-28.42 |  |
| **Imaging diagnosis** |  |  |  |
|  | Mean | 12.47 | 1.78 |
|  | CI95% | 11.18-13.76 | 1.31-2.24 |
| **Supplies** |  |  |  |
|  | Mean | 4.04 | 0.62 |
|  | CI95% | 2.90-5.19 | 0.29-0.95 |
| **Oxygen supply** |  |  |  |
|  | Mean | 43.05 | 0.07 |
|  | CI95% | 34.24-51.86 | 0.03-0.10 |
| **Overhead\*\*** |  |  |  |
|  | Mean | 8.87 | 10.70 |
|  | CI95% | 4.07-13.66 | - |
| **Equipment depreciation\*\*\*** |  |  |  |
|  | Mean | 1.33 | 0.17 |
|  | CI95% | 1.22-1.43 | 0.12-0.21 |
| **Total** |  |  |  |
|  |  | **Hospitalization** | **One visit to the pediatrician** |
|  | Median | 347.11 | 29.31 |
|  | Mean | 473.06 | 34.13 |
|  | CI95% | 399.16-546.97 | 32.60-35.66 |
| **Total (for one episode)\*\*\*\*** |  |  |  |
|  |  | **All visits+hospitalization** | **All visits** |
|  | Median | 425.40 | 84.12 |
|  | Mean | 528.32 | 85.92 |
|  | CI95% | 454.01-602.64 | 80.51-91.34 |

\* Patient and one parent (either food or milk for patients under 6 months old), only for hospitalized patients
\*\* Bedding and linens, maintenance and administrative expenses.
\*\*\*Includes equipment for mechanical ventilation, vital signs monitor, satsurometer, infusion pump, and diagnostic imaging equipment
\*\*\*\*Patients may have more than one visit to the pediatrician during a wheezing episode or before hospitalization

When we analyzed the cost by patient, the average cost for a hospitalization was US$544.88 (95% CI 370.67-719.09), whereas the cost for an ambulatory health visit was US$58.67 (95% CI 48.13-69.22). Finally, we calculated the cumulative direct medical cost per patient, encompassing both severe RSV-associated ALRTI and the total long-term wheezing episodes, resulting in a mean of US$859.44 (95% CI 739.52- 979.37)^26^.

Similarly, as in the RSV-associated ARLTI COI estimation, labor costs of healthcare workers was a major cost driver of the overall cost (Table 2), constituting between 40 to 76% of the total expenses^26^.

We have analyzed each direct medical cost variable, considering age groups across all episodes (Table 3), as well as variations in severity (Suppl. Table 1 and 2). We found that the total cost per episode of wheezing was significantly higher (p<0.001) in those infants younger than 12 months of age (207.43, 95%CI 154.3-260.6) compared with the subgroups of older toddlers. This difference can be partially explained by higher hospitalization rates in this age group, leading to increased expenses in laboratory examinations, health workers costs, and oxygen supply prices. No significant age-related differences were observed among children hospitalized due to severe obstructive wheezing exacerbations (Suppl. Table 1). In contrast, when evaluating the costs associated with outpatient episodes, it became evident that the older subgroup of patients, specifically those aged over 24 months, exhibited a markedly higher total cost (mean $77.35, 95%CI $65.62 - $89.08, p<0.001) along with significantly elevated costs across each comparative variable (Suppl. Table 2). Although fewer hospitalizations were recorded in children older than 2 years of age, the number of visits to the pediatrician, the severity of the wheezing episode and therefore the need of treatment in the emergency-room before discharge were higher (p<0.001).

**Table 3.** Comparison of costs of wheezing episodes by age group.

| Costs | Wheezing episodes after Severe RSV-ALRTI |  |  |  |  |
| --- | --- | --- | --- | --- | --- |
|  | Total episodes<br>N=429 | 0-12 months<br>(n=177) | 12-24 months<br>(n=152) | >24 months<br>(n=100) | p |
| **Laboratory cost** |  |  |  |  |  |
| Mean | 1.90 | 3.25 | 0.89 | 1.04 | **0.0004** |
| CI95% | 1.33-2.46 | 2.05-4.45 | 0.41-1.37 | 0.20-1.88 |  |
| **Staff cost** |  |  |  |  |  |
| Mean | 98.89 | 153.58 | 62.24 | 57.79 | **0.000** |
| CI95% | 79.02-118.76 | 111.43-195.72 | 42.86-81.62 | 33.85-81.74 |  |
| **Drug cost** |  |  |  |  |  |
| Mean | 8.31 | 8.31 | 7.51 | 9.54 | 0.072 |
| CI95% | 7.66-8.96 | 7.31-9.31 | 7.05-7.96 | 7.46-11.62 |  |
| **Feeding cost\*** |  |  |  |  |  |
| Mean | 5.32 | 7.42 | 3.35 | 4.60 | 0.153 |
| CI95% | 3.48-7.16 | 4.31-10.53 | 0.74-5.96 | 0.47-8.72 |  |
| **Imaging diagnostic cost** |  |  |  |  |  |
| Mean | 4.39 | 5.01 | 3.55 | 4.58 | 0.142 |
| CI95% | 3.75-5.04 | 3.85-6.17 | 2.67-4.43 | 3.31-5.85 |  |
| **Supplies cost** |  |  |  |  |  |
| Mean | 1.46 | 1.69 | 0.90 | 1.89 | 0.117 |
| CI95% | 1.06-1.85 | 1.01-2.37 | 0.40-1.39 | 0.94-2.84 |  |
| **Oxygen cost** |  |  |  |  |  |
| Mean | 10.59 | 17.46 | 6.43 | 4.74 | **0.0002** |
| CI95% | 7.83-13.35 | 11.57-23.35 | 3.37-9.49 | 2.26-7.22 |  |
| **Facilities cost** |  |  |  |  |  |
| Mean | 10.25 | 10.18 | 9.59 | 11.38 | 0.523 |
| CI95% | 9.09-11.41 | 8.36-12.00 | 8.96-10.23 | 7.65-15.11 |  |
| **Equipment cost** |  |  |  |  |  |
| Mean | 0.45 | 0.53 | 0.37 | 0.44 | 0.091 |
| CI95% | 0.39-0.51 | 0.42-0.64 | 0.28-0.46 | 0.31-0.57 |  |
| **Total cost** |  |  |  |  |  |
| Mean | 141.56 | 207.43 | 94.84 | 96.00 | **0.0001** |
| CI95% | 116.22-166.90 | 154.29-260.57 | 69.47-120.20 | 62.43-129.57 |  |

### Indirect costs and out of pocket expenses

Complete data on out-of-pocket expenses and indirect expenses were available for the families of 89 individuals (as shown in Suppl. Table 3). The average non-medical expenses associated with wheezing episodes in the cohort per individual was US$16.36 (95% CI $13.45 - $19.27), and per episode was US$ 4.89 (95% CI $4.27 - $5.50). The main driver of non-medical costs was food expenditures. Expenses were significantly higher in those hospitalized patients.

### Total cost related to long-term wheezing episodes

Per wheezing episode, the total cost including medical and non-medical expenses was US$ 191.01 (95% CI $166.37 - $215.64). Based on the level of care, we observed that the total cost for hospitalizations was significantly higher (US$535.55, 95%CI $461.25 - $609.84) compared to outpatient episodes (US$ 92.34, 95%CI $86.74 – $97.94). In table 4 we presented the cumulative cost per individual, encompassing all wheezing episodes with and without the inclusion of acute RSV hospitalizations. Interestingly, the average total cost of all the wheezing episodes per child was US$ 415.99 with a maximum cost of US$ 5391.96, while adding RSV hospitalization increase the cost to US$ 959.56 with a maximum of US$ 6024.35.

**Table 4.**
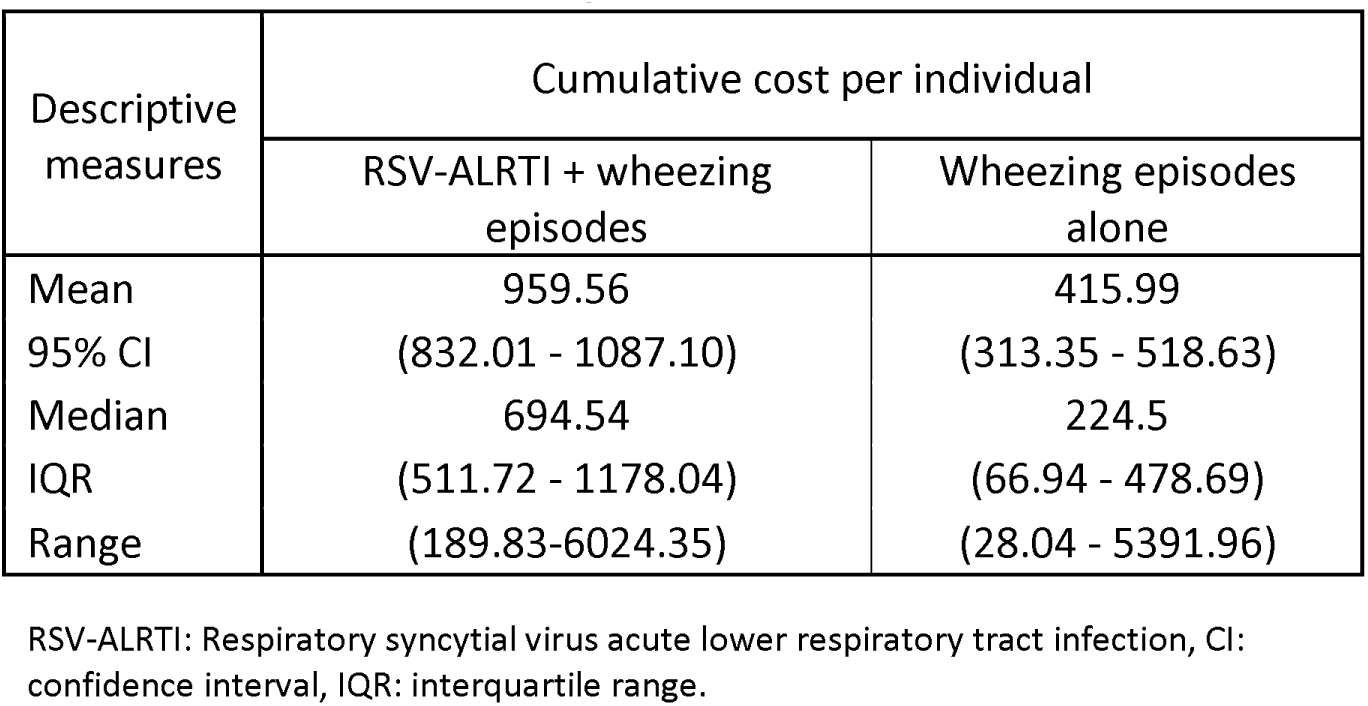
Costs accumulated by individuals.

## Discussion

This study explores the costs associated with long-term wheezing episodes after a severe RSV-ALRTI in the first year of life within a cohort treated at two public institutions in Buenos Aires, Argentina based on primary data. This research estimates the comprehensive economic impact of both RSV disease and their lasting effects. The findings underscore the substantial economic burden imposed by RSV in Argentina, emphasizing the significance of the study’s insights for future interventions against RSV globally. Upon analyzing the cumulative number of outpatient and hospitalized wheezing episodes, it was observed that in over fifty percent of patients who experienced severe RSV infection in their first year, the total costs were double that estimated for a single hospitalization due to ALRTI in public hospitals of Argentina^26^. However, the ranges of cost estimates can vary significantly, especially in patients with severe respiratory infections and frequent exacerbations leading to numerous hospitalizations, where costs can be up to 6 times higher than the average.

Overall, there is scarce information regarding the economic consequences of wheezing episodes after severe RSV-bronchiolitis, particularly in LMICs^13,27^. A recent study conducted in Colombia attempted to estimate the overall cost of RSV, accounting for long-term compications^18^. The authors reported a daily cost per patient with long-term complications of US$840.52 (SD 189.79). However authors estimated this cost based on a decision model rather than direct estimations as we did in this approach^18^. A retrospective study conducted in Japan highlighted that RSV produces a significant higher long-term economic burden when compared to other pathogens in both preterm and term children during their early years of life^44^. The authors demonstrated a higher cumulative health cost associated with severe RSV disease. Nevertheless, the cumulative costs associated with severe RSV disease and its long-term effects in Japan are notably higher than those estimated in our cohort^44^. Finally, the long-term utilization of healthcare resources in the United States following infant RSV infection had a substantial impact. This influence persisted for at least five years post-infection, with the majority occurring within the initial two years of life, aligning closely with the results obtained in this study^45^.

The healthcare system in Argentina is a mix of public, private, and social security providers^46^. It is decentralized, with each province managing its own healthcare services^46^. The public healthcare system provides services to the entire population, while those with private insurance have access to a network of private clinics and hospitals^46–48^. Challenges include disparities in healthcare quality between urban and rural areas and variations in access to specialized services^8,49^. Argentina follows a vaccination policy that includes a comprehensive National Immunization Program, that provides free vaccines to the entire population, aiming to prevent a range of infectious diseases^50^. In Argentina RSV prevention policy is relevant. In this sense, the use of palivizumab was implemented several years ago and its use recommendation is guided by national health authorities for specific subgroup of risk infants^51^. Argentina has implemented diverse public health policies aimed at enhancing health outcomes and mitigating overall healthcare costs. These initiatives include vaccination campaigns, maternal and child health programs, screenings for common diseases, promotion of primary healthcare services, advocacy for the use of generic medications, and the implementation of the Remediar plan, ensuring free and equitable access to essential medications for individuals and communities with limited resources^46,52^. Finally, for forthcoming public health decisions, it is vital that the Argentine government acquires accurate information on both the economic and epidemiological burdens of diseases, as outlined in this study. This data is indispensable for a comprehensive evaluation of health technologies. However, this study was conducted at two public hospitals situated in the southern metropolitan area of Buenos Aires. Hence, there is a need to broaden the investigation of costs, considering the perspectives of private and social security providers, which were not addressed in this study.

Our study has some limitations. Primarily, as the economic analysis was integrated into a more extensive prospective study, acquiring detailed and continuous information regarding families’ out-of-pocket and indirect expenses was not feasible. Furthermore, considering the prevalence of high unemployment and informal work in a population characterized by very low socioeconomic status, there is a potential underestimation of family expenses. To mitigate this challenge, we have applied a two-part model to address the substantial proportion of zero observations combined with biased positive results^38,39^. However, even with these efforts, we believe our estimates of household expenses may still be underestimated. Lastly, it’s essential to note that the Argentine government has introduced various subsidies for medications, supplies, and general expenses, potentially resulting in an reduction of the costs presented in this study^52^. To gain a comprehensive understanding, it is imperative to analyze costs from private healthcare perspectives and consider variations across different regions of the country.

The recent endorsement of preventive interventions against RSV by the U.S. Food and Drug Administration and the European Medicines Agency signifies a transformative shift in the paradigm of RSV disease^53^. Various countries’ government authorities now face the crucial task of assessing the feasibility of incorporating these preventive measures into routine medical practices, giving rise to diverse implementation scenarios^53^. Hence, this study holds significant relevance, as the information it generates could allow us to understand that severe RSV disease has a short, medium and long-term impact not only from a clinical but also an economic perspective. The Argentine Ministry of Health has recently approved for implementation the maternal vaccine against RSV paving the way for a change in the burden of disease^54^. Consequently, these insights will prove valuable in shaping future cost-effectiveness models.

Our study aligns with the objective of tracking infants until the age of five. Specifically, we investigated the long-term economic burden of wheezing associated with RSV in Argentine infants diagnosed before this age. The findings indicate that preventing RSV in the first year of life could significantly influence older children’s health and economic outcomes.

## Data Availability

The datasets used and/or analyzed during the current study are available in Zenodo (https://zenodo.org/records/10440099) or from the corresponding author.

## List of abbreviations

RSV: respiratory syncytial virus
ALRTI: acute lower respiratory tract infection
CI: confidence interval
COI: cost of illness
ER: emergency room
GLM: generalized linear model
IQR: interquartile range
LMICs: low- and middle-income countries
OLS: ordinary least squares
SD: standard deviations

## Ethics approval and consent to participate

This study, observational in design, used retrospective data, and was conducted in accordance with the amended Helsinki Declaration, the International Guidelines for Ethical Review of Epidemiological Studies, and Argentinean laws on data protection and patients’ rights. This study implies the use of pseudonymized individual data, using double dissociation (i.e., in the original data source and once data are stored in the database) which impedes patients re identification. The institutional review boards at each participating hospital in the Province of Buenos Aires approved the study (approval ID numbers: 025/21 and 028/21). Informed consent was obtained from all participating parents or guardians.

## Patient and Public Involvement

Patients or the public WERE NOT involved in the design, or conduct, or reporting, or dissemination plans of our research.

## Availability of data and materials

The datasets used and/or analyzed during the current study are available from the corresponding author on reasonable request.

## Authors’ contributions

RL, MTC, FPP, and CP conceptualized and designed the study. MTC and RL were directly involved in the post validation of the results. RL, ES, JD, and MTC created all the data capture forms. KD, AS, ES, and PC reviewed all the instruments and obtained all the primary information. JD, ES, and MTC provided data management tasks and quality assessment of data previous to run the analysis. JD, CRR, and MTC were in charge of the methodology and analyzed the data. JD and MTC prepared the original draft. MTC and CP supervised the whole process. All authors reviewed, edited, and approved the final manuscript for submission and accept public access. PATH were involved in the study design, data collection, data analysis, data interpretation, and writing of the manuscript. All authors had full access to all the data in the study, and the corresponding author had final responsibility for the decision to submit for publication.

## Acknowledgements

Authors would also like to extend their thank to Adrian Ferretti the laboratory technician and the regulatory and administrative staff of Fundación Infant (Alejandra and Veronica Bianchi, Carola Candurra, Ana Ramos Aloi, and Roxana Olivera) for their support.

