## Supplementary material for "Assessing the Long-Term Economic Impact of Wheezing Episodes After Severe RSV Disease in Children from Argentina: A Cost of Illness Analysis"

Affiliations: 1 Fundación Infant, Buenos Aires, Argentina, 2 Escuela de Bio y Nanotecnologías, Universidad Nacional de San Martín (UNSAM), San Martín, Provincia de Buenos Aires, Argentina, 3 Consejo Nacional de Investigaciones Científicas y Técnicas (CONICET), Argentina, 4 Center for Vaccine Innovation and Access, PATH, Seattle, Washington, USA, 5 Servicio de Pediatría, Hospital Evita de Lanús, Provincia de Buenos Aires, Argentina, 6 Servicio de Pediatría, Hospital Evita Pueblo de Berazategui, Provincia de Buenos Aires, Argentina. 7 Centre for Health Economics, University of York, Heslington, York, United Kingdom. 8 Facultad de Ciencias Económicas, Universidad Nacional de La Plata, Argentina.

**Study procedures**

The study evaluated costs associated with ALRTIs and associated wheeze among children identified as RSV+ in Buenos Aires. The costing study approach included 3 main components: (1) evaluation of resources utilized during care for ALRI and wheeze among children with RSV; (2) assessment of direct medical costs for each type of resource identified; (3) identification of non-medical and indirect costs associated with seeking care.

(1) We assessed healthcare resource utilization associated with acute ALRI and subsequent wheezing episodes. Resource utilization includes medical procedures, bed nights, tests, medications, and other components of care. We will assess resource utilization via secondary data analysis of the existing study database. The database contains information on healthcare resource utilization, RSV diagnosis, and demographic information from patients experiencing ALRIs and subsequent wheeze. Healthcare utilization data will be pulled for each child to identify resources used during the child’s course of treatment in the facility. If any data gaps are identified during the secondary analysis, we conducted a medical record review for any enrolled children for whom data is not currently available through the study dataset.
(2) We collected direct medical costs of each resource identified for treatment of ALRI and wheeze. Direct medical cost data to be collected include medical procedures, medications, laboratory tests, consumables, and bed nights, among others. These costs, as well as non-medical overhead costs, were obtained from hospital administrators, financial officers, pharmacy administrators, supply chain managers, logisticians/procurement staff, laboratory managers, and others with knowledge about hospital finances and/or itemized unit costs. Detailed cost data are important to calculate the full cost to the healthcare system and important for cost-effectiveness evaluation from government provider perspective.
(3) **For costs of long-term wheeze only:** Patient-level out-of-pocket, non-medical (e.g., transportation), and indirect costs (e.g., opportunity costs of time missed from work) incurred during wheeze episodes were obtained via phone interviews with parents/guardians of children who were enrolled in the larger study. Prior to conducting the interviews, we re-consented parents/guardians via phone for their participation in the costing activities. Ongoing data collection was underway by the study staff to prospectively capture information about wheeze episodes that occur after the ALRI event. Parents/guardians of children with wheeze were quarterly called by researchers at to collect information on wheeze events that occurred since the previous call. We will add economic questions to the current questionnaires used by the study to assess out-of-pocket, non-medical and indirect costs. In addition to costs of seeking care, households will be asked about post-discharge costs, household assets, income, and sociodemographic factors. Questions were also included to identify any expenditures foregone by the household due to medical related costs, i.e. food, school fees, etc. Participants were asked if they have had additional facility visits for wheeze since the prior follow-up call. Costs for any related household visits were also collected. These data allowed us to determine household costs associated with the child’s illness, information that is critical for calculating the full societal cost of illness potentially avertable by vaccination. The follow-up calls were conducted by a trained researcher and were conducted over the course of one year. All parents/guardians were consented for their participation in answering the economic questions added to current questionnaires.
(4) Healthcare utilization and cost data were combined at the individual patient level. For each patient we enroll in the costing study, we estimated the total cost of treatment by multiplying the health care resources utilized by the costs of each resource and calculating the sum of products.

Data was collected by trained researchers and/or study coordinators. Confirmation of historical RSV was obtained through the existing database by a researcher or clinical team member. All clinical, resource utilization and cost data were entered into a stand-alone database for the cost of illness analysis. Clinical data was validated by review of 30% of patient records by a second researcher. Quality of data entry for questionnaire responses will be conducted by comparing quality checks on 30% of survey data entries. Questionnaires that have missing data were reviewed to determine the reason why the data is missing. If the coordinator determined that it was possible to recover the data, study staff attempted to obtain the missing information. Double data entry was conducted for a proportion of questionnaires as a supplemental quality control measure. Data validation and data cleaning occurred in real-time throughout data collection period.

Enrollment

2014

2015

2017

2016

2019

2018

2020

2021

2022

qPCR measure

COI-analysis

2023

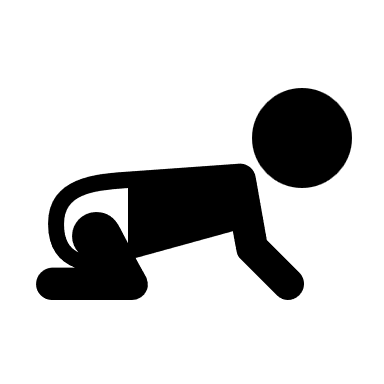

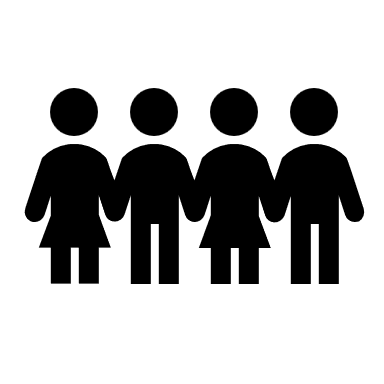

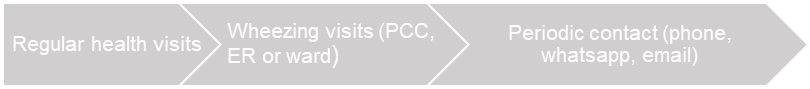

Follow-up for wheezing diagnosis

Regular health visits

Wheezing visits (PCC, ER or ward)

Periodic contact (phone, whatsapp, email)

**Supplementary Figure 1. Flow chart of the study procedures.**

| **Supplementary table 1. Comparison of costs of wheezing episodes by age group in hospitalized patients** | | | | | |
| --- | --- | --- | --- | --- | --- |
| **Costs** | **Wheezing episodes in hospitalized patients** | | | | |
|  | **Total episodes** | **0-12 months** | **12-24 months** | **>24 months** | **p** |
|  | **N=105** | **(n=62)** | **(n=28)** | **(n=15)** |  |
| **Laboratory cost** |  |  |  |  |  |
| Mean | 7.58 | 8.98 | 4.84 | 6.92 | 0.189 |
| CI95% | 5.64-9.53 | 6.03-11.93 | 2.71-6.96 | 1.88-11.97 |  |
| **Staff cost** |  |  |  |  |  |
| Mean | 361.33 | 414.34 | 279.20 | 295.53 | 0.087 |
| CI95% | 303.88-418.78 | 324.02-504.65 | 221.95-336.45 | 198.02-393.04 |  |
| **Drug cost** |  |  |  |  |  |
| Mean | 12.66 | 12.19 | 10.88 | 17.93 | 0.142 |
| CI95% | 10.43-14.89 | 9.59-14.78 | 9.00-12.75 | 6.17-29.69 |  |
| **Feeding cost*** |  |  |  |  |  |
| Mean | 21.74 | 21.18 | 18.20 | 30.65 | 0.525 |
| CI95% | 15.05-28.42 | 13.26-29.10 | 4.77-31.64 | 4.69-56.61 |  |
| **Imaging diagnostic cost** |  |  |  |  |  |
| Mean | 12.47 | 12.56 | 11.57 | 13.73 | 0.593 |
| CI95% | 11.18-13.76 | 10.47-14.66 | 10.69-12.45 | 11.20-16.27 |  |
| **Supplies cost** |  |  |  |  |  |
| Mean | 4.04 | 4.33 | 3.23 | 4.37 | 0.702 |
| CI95% | 2.90-5.19 | 2.67-5.99 | 1.46-5.01 | 1.28-7.46 |  |
| **Oxygen cost** |  |  |  |  |  |
| Mean | 43.05 | 49.79 | 34.73 | 30.73 | 0.184 |
| CI95% | 34.24-51.86 | 36.10-63.49 | 22.32-47.14 | 21.93-39.53 |  |
| **Facilities cost** |  |  |  |  |  |
| Mean | 8.87 | 9.21 | 4.70 | 2.39 | 0.325 |
| CI95% | 4.07-13.66 | 3.92-14.50 | 1.75-7.64 | 1.84-2.95 |  |
| **Equipment cost** |  |  |  |  |  |
| Mean | 1.33 | 1.35 | 1.25 | 1.39 | 0.668 |
| CI95% | 1.22-1.43 | 1.20-1.49 | 1.12-1.38 | 0.99-1.78 |  |
| **Total cost** |  |  |  |  |  |
| Mean | 473.06 | 533.92 | 368.60 | 416.51 | 0.135 |
| CI95% | 399.16-546.97 | 419.41-648.44 | 286.97-450.24 | 269.60-563.41 |  |

| **Supplementary table 2. Comparison of costs of wheezing episodes by age group in outpatients** | | | | | |
| --- | --- | --- | --- | --- | --- |
| **Costs (mean, SD)** | **Wheezing episodes in outpatients** | | | | |
|  | **Total episodes** | **0-12 months** | **12-24 months** | **>24 months** | **p** |
|  | **N=324** | **(n=115)** | **(n=124)** | **(n=85)** |  |
| **Laboratory cost*** |  |  |  |  |  |
| Mean | - | - | - | - | - |
| CI95% |  |  |  |  |  |
| **Staff cost** |  |  |  |  |  |
| Mean | 13.84 | 13.00 | 13.25 | 15.84 | **0.012** |
| CI95% | 13.04-14.64 | 12.10-13.90 | 12.23-14.27 | 13.49-18.19 |  |
| **Drug cost** |  |  |  |  |  |
| Mean | 6.90 | 6.22 | 6.74 | 8.06 | **0.0007** |
| CI95% | 6.53-7.28 | 6.03-6.41 | 6.51-6.98 | 6.70-9.42 |  |
| **Imaging diagnostic cost** |  |  |  |  |  |
| Mean | 1.78 | 0.94 | 1.74 | 2.96 | **0.004** |
| CI95% | 1.31-2.24 | 0.34-1.54 | 0.99-2.50 | 1.84-4.09 |  |
| **Supplies cost** |  |  |  |  |  |
| Mean | 0.62 | 0.27 | 0.37 | 1.45 | **0.012** |
| CI95% | 0.29-0.95 | 0-0.64 | 0-0.79 | 0.47-2.42 |  |
| **Oxygen cost** |  |  |  |  |  |
| Mean | 0.07 | 0.03 | 0.04 | 0.16 | **0.012** |
| CI95% | 0.03-0.10 | 0-0.08 | 0-0.09 | 0.05-0.26 |  |
| **Equipment cost** |  |  |  |  |  |
| Mean | 0.17 | 0.09 | 0.17 | 0.27 | **0.004** |
| CI95% | 0.12-0.21 | 0.03-0.14 | 0.10-0.24 | 0.17-0.37 |  |
| **Total cost (one visit)** |  |  |  |  |  |
| Mean | 34.13 | 31.40 | 33.02 | 39.44 | **0.0001** |
| CI95% | 32.60-35.66 | 29.65-33.15 | 31.13-34.90 | 34.99-43.89 |  |
| **Total cost (all visits)** |  |  |  |  |  |
| Mean | 58.96 | 48.27 | 56.26 | 77.35 | **0.000** |
| CI95% | 54.42-63.50 | 43.18-53.37 | 49.46-63.06 | 65.62-89.08 |  |

| **Supplementary table 3. Non-medical expenses.** | |
| --- | --- |
| **Transport** | **n=109** |
| Median | 4.25 |
| Mean | 4.54 |
| CI 95% | 4.12-4.97 |
| **Food** | **n=119** |
| Median | 5.52 |
| Mean | 7.85 |
| CI 95% | 6.34-9.36 |
| **Indirect expenses** | **n=146** |
| Median | 2.53 |
| Mean | 4.14 |
| CI 95% | 3.36-4.93 |
| **Total** | **n=89** |
| Median | 12.08 |
| Mean | 16.36 |
| CI 95% | 13.45-19.27 |
